# Malaria awareness of adults in high, moderate and low transmission settings: A cross-sectional study in rural East Nusa Tenggara Province, Indonesia

**DOI:** 10.1101/2020.12.04.20243675

**Authors:** Robertus Dole Guntur, Jonathan Kingsley, Fakir M. Amirul Islam

**Author notes:** Corresponding author: (RDG). Department of Health Science and Biostatistics, Swinburne University of Technology, Hawthorn Campus, Victoria, Australia 3122 (Robertus Dole Guntur, Fakir M. Amirul Islam). Department of Health and Medical Science, Swinburne University of Technology, Hawthorn Campus, Victoria, Australia 3122 (Jonathan Kingsley).

## Abstract

**Introduction:** The Indonesian roadmap to malaria elimination in 2009 indicated that the nation is progressing towards achieve malaria elimination by 2030. Currently, most of the districts in the Western part of Indonesia have eliminated malaria, however, none of the districts in East Nusa Tenggara Province (ENTP) have met set targets. This study aims to investigate the status of malaria awareness of rural adults in the ENTP.

**Methods:** A cross-sectional study was conducted between October and December 2019 in high, moderate, and low malaria endemic settings (MES) in ENTP. 1495 participants recruited by multi-stage sampling method were interviewed using a validated questionnaire, after obtaining informed consent. A malaria awareness index was developed based on ten questions. Logistic regression method was applied to investigate the significance of associations of malaria awareness with the three malaria endemic settings.

**Results:** Participants were between the age of 18 and 89 years old, 51.4% were female and 45.5% had completed primary education. Malaria awareness index was very low (48.8%, 95% confidence interval (CI): 45.2 – 52.4). Malaria awareness of rural adults residing in low endemic settings was three times higher compared to those were living in high endemic settings (Odds ratio (OR): 3.11, 95% CI: 2.40 – 4.03, p < 0.001) and the basic malaria knowledge for participants living in low malaria endemic setting was almost five times higher than that of in high endemic setting (OR: 4.66, 95% CI: 3.50 – 6.20, p < 0.001). Of total participants, 81.3% (95% CI: 79.1 – 83.5) were aware that malaria could be prevented and 75.1% (95% CI: 72.6 – 77.6) knew at least one prevention measure. Overall, the awareness of fever as the main symptom of malaria, mosquito bites as the transmission mode of malaria, and seeking treatment within 24 hours when suffering with malaria was poor, 37.9% (95% CI: 33.9 – 41.9), 59.1% (95% CI: 55.9 – 62.3), and 46.0% (95% CI: 42.3 – 49.7) respectively. The poor level of awareness was statistically significantly different amongst three MES, the level of awareness was the lowest in the high endemic setting.

**Conclusion:** Malaria awareness of rural adults needs to be improved to address Indonesia’s national roadmap to malaria elimination. Results indicate public health programs at a local government level should incorporate the malaria awareness index in their key strategic intervention packages to address local malaria awareness.

## Introduction

Malaria is a major global health problem with an estimated 1.2 billion people living at high risk of being infected [1]. However, malaria cases and associated death have decreased in the last decade [2]. From 2010 to 2018, the total number of malaria cases decreased by about 1% per year and deaths due to malaria declined by 5% per year [1]. The number of countries reporting that local transmission less than 51 cases increased from 5 countries in 2010 to 11 countries in 2018 [3].

Countries having zero local transmission in the last three consecutive years are eligible for requesting malaria elimination certification from WHO [3]. In South-East Asia Region (SEARO), two countries, the Maldives and Sri Lanka have been certified malaria-free areas by the WHO [4]. In alignment with the global action plan for a malaria-free world [5] and global technical strategy for malaria elimination [6], the WHO SEARO action plan indicates that all countries in the region will be malaria-free zones by 2030 [4].

The road map of malaria elimination in Indonesia has been declared since April 2009, proclaiming that Indonesia also aims to eliminate malaria by 2030 [7, 8]. All malaria endemic districts in Indonesia were divided into four categories based on the annual prevalence incidence (API) [9]. Currently, 55.5% of the total number of districts in the country (285 out of 514 districts) has been categorized as malaria elimination districts in 2018 [9]. All of the districts in the provinces of DKI Jakarta, Bali and East Java have been categorized as malaria elimination areas, whereas none of the districts from five provinces in the eastern part of Indonesia such as Papua, West Papua, Maluku, North Maluku, and East Nusa Tenggara Province (ENTP) has achieved this categorization [9].

The ENTP is a province with an API value five times higher compared to the national API of Indonesia [10]. This province has 21 districts and one municipality [11]. Ten districts and a municipality are low endemic meanwhile the number of districts has been classified as moderate and high endemic is six and five districts respectively [9]. In line with the national commitment to eliminate malaria by 2030, various effort of the local authority have been done to support malaria elimination in this province. This includes increasing coverage of artemisinin-based combination therapy (ACT) as the first line of malaria treatment from 55% in 2013[12] to 83.1% in 2018 [10] and screening malaria for pregnant women during her first visiting to local health centres [13]. For controlling mosquitoes, the introduction of treated bed net has been implemented in the most of the districts in the region since 2008 [14], mass distribution of the long-lasting insecticide-treated nets (LLINs) for 15 districts in 2017 [15], and the use of special repellent [16]. However, the number of malaria cases are still high (17,192 cases) [17] indicating that those interventions might be ineffective and the implementation of those interventions might depend on the community behaviour which was limited investigated in this province. In fact, knowledge and behaviour of the community play a significant role in supporting malaria elimination [18, 19]. Having high level of malaria awareness in communities enables them to improve self-protection [20], seeking early treatment [21], reducing malaria prevalence [22], and consequently speeding up malaria elimination [23].

A number of studies around malaria knowledge have been undertaken in Indonesia since the declaration of national commitment to eliminate malaria [24-27]. Knowledge on LLINs, which is the most effective tool to prevent malaria [28] and currently adopted as the primary vector control interventions in many parts of Indonesia [8], was not investigated in those studies. Also, most of the studies were conducted in the western part of the country, most of which have been classified as malaria elimination district. Studies were also conducted at sub-district and village levels. One population-based study on 4,050 participants in North Maluku Province revealed that only about half of the respondents knew about symptoms of malaria and the majority of participants (98%) did not know the main cause of malaria [27]. However, approximately 50% of the participants were less than 18 years old which was hardly suitable candidates for measuring the level of knowledge of a particular community.

In the ENTP, various studies of malaria knowledge were conducted [13, 29-31]. Most of these studies were conducted at village and subdistrict level. One population study covering only pregnant women in high malaria endemic area of the province indicating that low level of malaria prevention knowledge particularly related to LLINs [13]. Another population level study related to community behaviour of malaria was conducted in ENTP in 2018 [31], however, the study investigated only the practice of malaria prevention of the rural community and the comparison of malaria prevention awareness amongst MES has not been investigated yet. To date, the investigation on malaria knowledge related to symptoms, transmission mode, prevention method and the perception of malaria treatment seeking behaviour of rural adults in different types of malarial endemic settings in the ENTP has not been performed yet. Investigation of malaria awareness of rural communities is critical for Indonesia considering that 52% of malaria cases in the country were contributed by rural communities [10] and variation of malaria prevention practice amongst provinces exists in the country [31]. Understanding the level of malaria knowledge of rural communities and finding which MES is most vulnerable is essential for the development and implementation of evidence-based strategies to accelerate progress for malaria elimination in the province. This research will fill this gap with the aim of investigating malaria awareness of rural adults in three different MES in order to support national commitment of Indonesia government to eliminate malaria by 2030.

## Materials and methods

### Study Population

East Nusa Tenggara Province (ENTP) is one of 34 provinces in Indonesia. It is in the eastern part of the country. The total population of ENTP is 5.3 million, contributing about 2.04% of the total population of Indonesia [11]. The ratio of male to female (50.5% to 49.5%) was comparable with that of Indonesia (50.2% to 49.8%). The area of the province is 47,931.54 km^2^, with the population density 114 people per square kilometre, located between 1180° and 1250° east longitudes and between 80° and 120° south latitudes [11]. This project was conducted in three regencies of different malaria endemic settings (MES) which are East Sumba, Belu, and East Manggarai district representing as high, moderate, and low MES respectively [32] as shown in S1 Fig 1.

**Fig 1.**
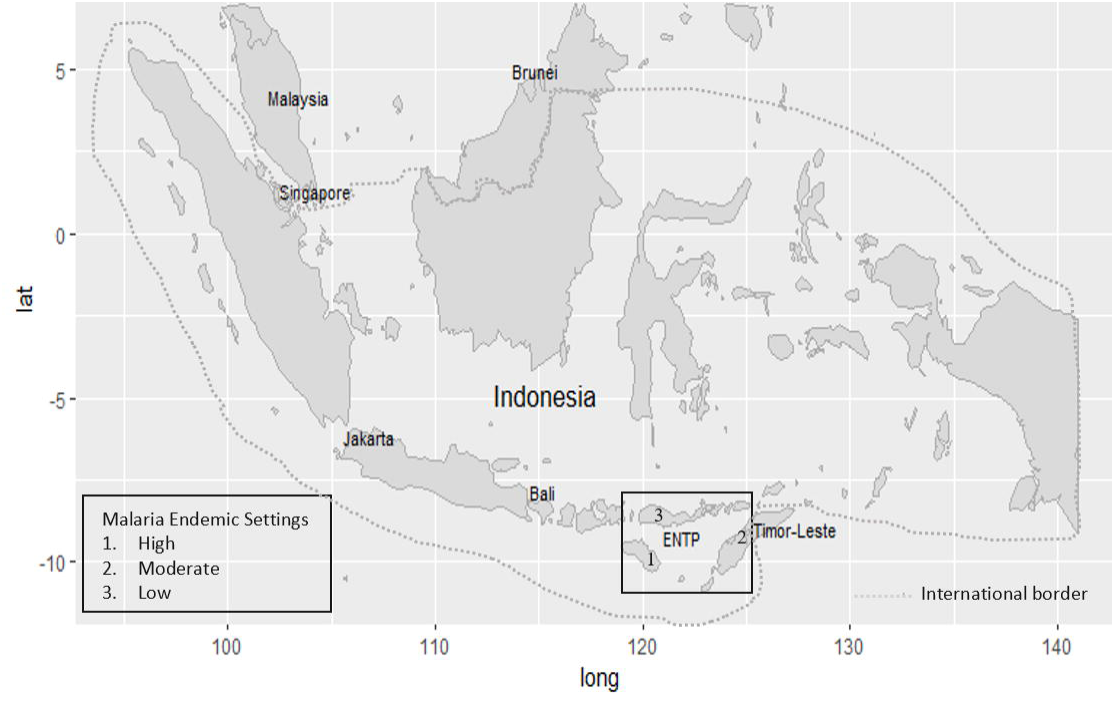
Map of Study Sites.

The sample was comprised of 1495 participants (51.4%, n = 768 female) aged from 18 to 89 recruited using multi-stage cluster sampling procedure with a systematic random sampling procedure at cluster level 4 from each of the total 49 villages from ENTP. The sample size was based on the prevalence of malaria in ENTP of 1.99% in 2018 estimated by the Indonesian basic health research, which is called Riskesdas [10]. Sample size calculation was described comprehensively in the previous publication of the authors [33]. Overall, the sample size was sufficiently large enough to detect a minimum 5% difference in the proportion of malaria awareness amongst high, moderate, and low malaria endemic settings (Statistical power > 90%, p=0.05).

### Recruitment strategy

A multi-stage cluster sampling procedure was applied to recruit adults from three districts. Three out of 22 districts have been selected based on the annual parasite incidence (API) of malaria. In each selected district, three sub-districts were randomly chosen. The number of clusters was selected from each sub-district based on their relative population. In each village, a systematic random sampling technique was used to get 20-40 participants per village proportionate to the population size of the villages. In the selected households, the research teams first approached the heads of the households for interviews. In case the household heads, either husband or wife, are absent, any residents over 18 years of age could serve as study participants [34].

### Data collection

The collection of data for this study was done in collaboration with local nurses who are local residence in the study area. Data collectors interviewed participants face-to-face based on the guidance of the structure questionnaire. This instrument was adapted from validated questionnaire that had been published by other authors [35, 36]. The English version of the questionnaire had been translated into the local language by the author of the article in the cooperation with local language experts. Furthermore, the translation version of the questionnaire was demonstrated upon 30 participants before finalising the questionnaire. On the first part of the questionnaire, we collected data on the socio-demographic variables of participants and the second part is questions used to investigate the understanding of general malaria knowledge of participants as shown in the questionnaire that has been published on the previous publication of the authors [33].

## Malaria awareness measure

Ten questions were used to assess malaria awareness of rural adults. The first three questions categorized as basic understanding of malaria including whether participants have heard malaria term, whether malaria was dangerous for their health, and whether malaria can be prevented. The response for these questions was yes or no. For participants answering yes, they got score one. Overall, participants obtained score three if they can answer correctly all these three questions. Total mark for the basic understanding of malaria was evaluated following the previous studies [37, 38]. Participants who can answer correctly at least 60% for the first three questions were categorized as having awareness in basic malaria understanding, otherwise categorized as having no awareness in basic malaria understanding.

The next seven questions categorized as basic malaria knowledge comprising whether participants could identify the main symptom and the main cause of malaria, whether participants could identify some protective measures to prevent malaria, whether participants seeking treatment for their malaria within 24 hours after the onset of the symptoms. Participants who could identify fever as the main symptom of malaria [39] and mosquito bites as the main cause of malaria [40, 41] obtained score one respectively. Participants who could mention sleeping under non-LLINs, sleeping under LLINs, using mosquito coils, keeping house clean as the method to prevent malaria achieved a score of one respectively. Finally, participants who mention seeking malaria treatment within 24 hours for their malaria [42] obtained a score of one. Each participant accomplished a total score of seven if they could answer correctly all seven questions. Total mark for the basic malaria knowledge was further evaluated following the procedure in the previous studies [37, 38]. Participants answering correctly at least 60% for these seven questions were categorized as having awareness in basic malaria knowledge.

Overall, each participant gained a total score of ten if they could answer correctly all these ten questions. Total marks for ten questions were evaluated following the previous studies [37, 38]. The level of malaria knowledge of participants with the accurate rate above 80%, 60 –79%, 1 – 59%, 0 were classified as excellent, good, poor, and zero, respectively. The participants who were in the group of excellent and good were categorised as having malaria awareness while those who were in the poor and zero group were classified as unaware of malaria [37, 38].

### Socio-demographic covariates

Demographic information comprising gender, age, education level, and socio-economic status (SES) was collected. Gender was categorised as male and female. Age was classified as 5 groups, less than 30 years old, 30 – 40 years, 40 – 50 years, 50 – 60 years and above 60 years old. The level of education was categorised as no education, primary school level of education (grade 1 to 6), junior high school (grade 7 to 9), senior high school (grade 10 to 12), and diploma or above education level. The SES group was assessed according to the ownership of durable asset and housing characteristics [43]. In each selected household, the participant was asked for ownership of ten items of durable assets including radio, television, electricity, bike, motorcycle, hand phone, fridge, tractor, generator and car. Housing characteristics were evaluated by having access to water taps in dwellings and main material of house. Houses having floor and wall constructed of cement were categorised as modern houses, otherwise as non-modern houses. In total, there are 12 items used to construct the SES level. Three levels of SES were defined by counting ownerships of the items. Low SES was defined as having zero or one item; moderate SES was defined as owning 2 – 4 items and high SES was defined as having more than 4 items as highlighted by Zafar et al. [43].

### Statistical Analyses

The participants’ socio demographic characteristics including gender, age group, education level, and SES were reported using description statistics. The association between malaria endemic settings and 10 questions was explored by chi-square method. This approach was further applied to evaluate initially the association of basic malaria understanding, basic malaria knowledge, the level of malaria knowledge, and the level of malaria awareness amongst three types of malaria endemic settings. To evaluate the strength of the association between different types of malaria endemic settings and basic malaria understanding, basic malaria knowledge, as well as malaria awareness, logistic regression model was applied. The P value of 0.05 or less was termed to be statistically significant. Statistical software SPSS version 27 (SPSS Inc.) was used for analyses.

### Ethics approval

The research was conducted in accordance with the tenet of The Declaration of Helsinki and was approved by Human Ethics Committee of the Swinburne University of Technology (Reference: 20191428-1490) and from Health Research Ethics Committee, National Institute of Health Research and Development (HERC-NIHRD) the Indonesian Ministry of Health (Reference: LB.02.01/2/KE.418/2019). Written consent was obtained from participants having full capacity to give voluntary consent in his/her own right on the basis of sufficient information provided. For participants who were unable to read the consent documentation, the participants gave their authority to their spouse or immediate family to read the consent form. Their spouse/immediate family provided a signature on the consent form on behalf of the participants and the ethics committee approved this consent procedure. Participants were informed of their right to withdraw from study at any stage or to restrict their data from their analysis.

## Results

### Demographic characteristics of the study population

Of the total participants aged between 18 and 89 years (mean 43.8 years and standard deviation 12.8 year), 51.4% was female. In terms of educational attainment, the majority of respondents had completed primary education (45.4%) and almost 20% of them did not have any formal education. The disparity of education distribution amongst these three settings was evidence, having no education in high MES was 35% compared to 2.6% in the low MES. Socio economic status of participants depicted that the majority of them (57.5%) living in moderate SES. The socio-demographic characteristics of the participants based on the malaria endemic setting are shown in Table 1.

**Table 1.**
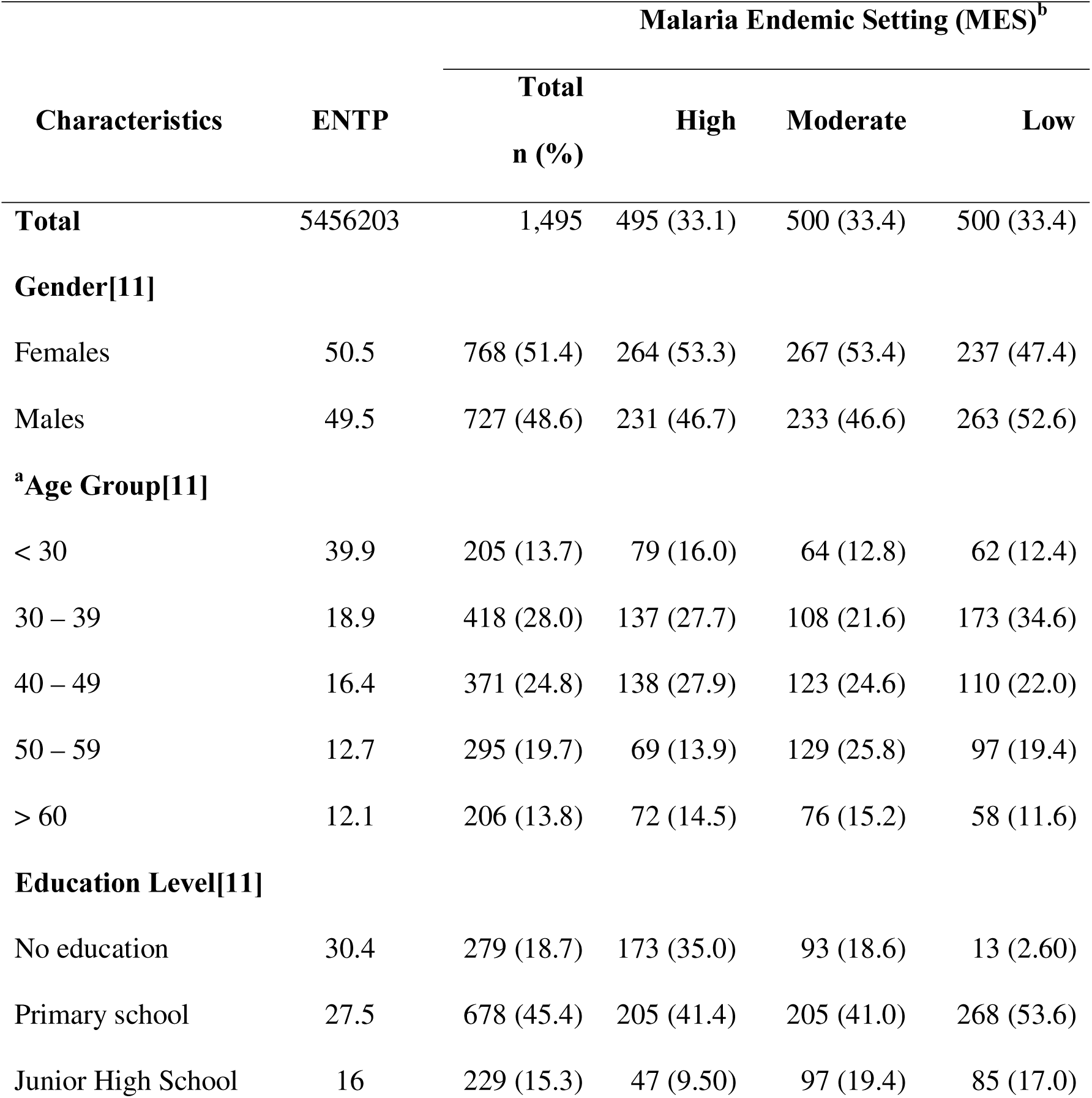
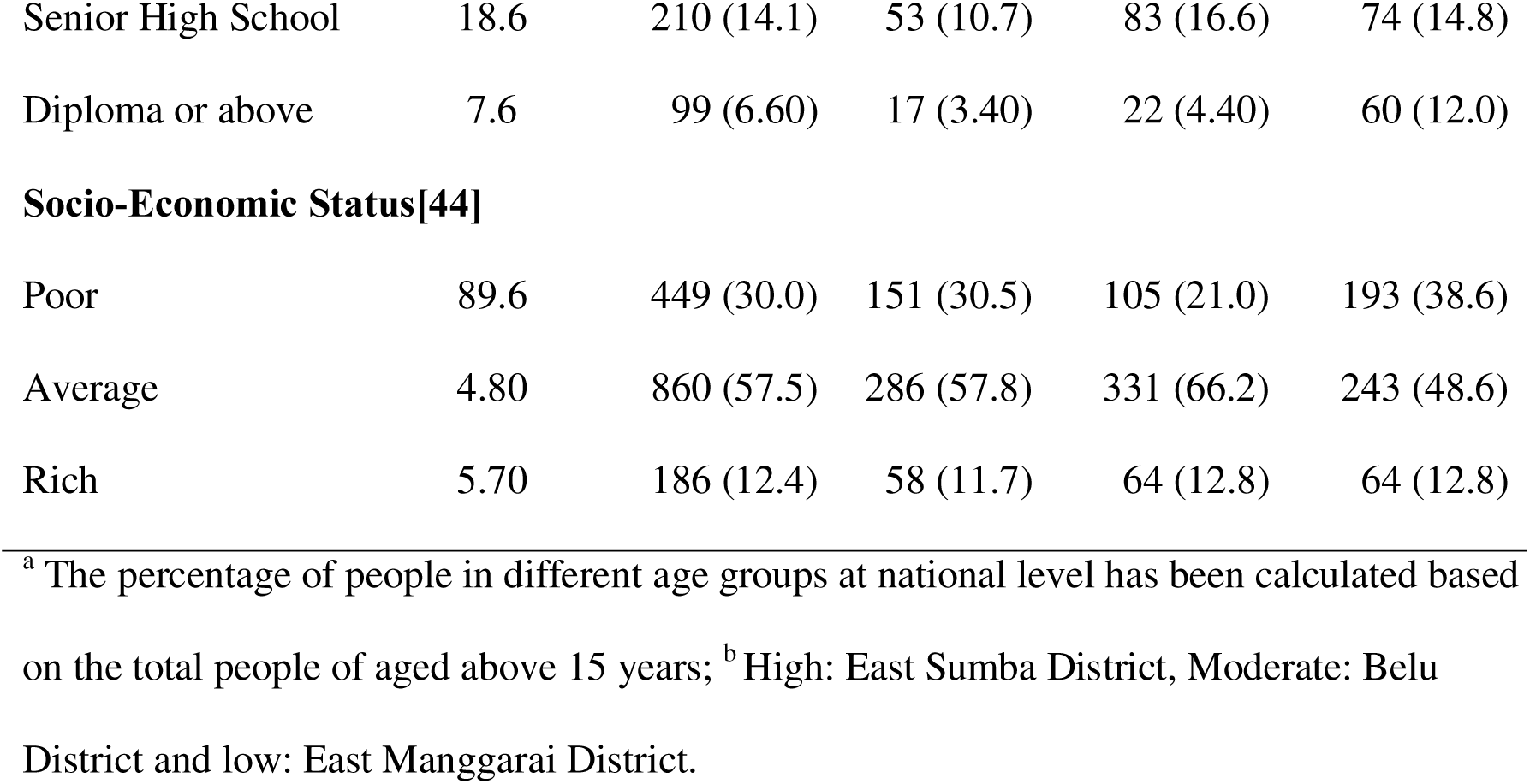
Distribution of study participants and participants from a national representative sample in three different MES in the East Nusa Tenggara Province (ENTP), Indonesia.

| Characteristics | ENTP | Malaria Endemic Setting (MES)<sup>b</sup> |  |  |  |
| --- | --- | --- | --- | --- | --- |
|  |  | Total<br>n (%) | High | Moderate | Low |
| **Total** | 5456203 | 1,495 | 495 (33.1) | 500 (33.4) | 500 (33.4) |
| **Gender[11]** |  |  |  |  |  |
| Females | 50.5 | 768 (51.4) | 264 (53.3) | 267 (53.4) | 237 (47.4) |
| Males | 49.5 | 727 (48.6) | 231 (46.7) | 233 (46.6) | 263 (52.6) |
| **<sup>a</sup>Age Group[11]** |  |  |  |  |  |
| < 30 | 39.9 | 205 (13.7) | 79 (16.0) | 64 (12.8) | 62 (12.4) |
| 30 – 39 | 18.9 | 418 (28.0) | 137 (27.7) | 108 (21.6) | 173 (34.6) |
| 40 – 49 | 16.4 | 371 (24.8) | 138 (27.9) | 123 (24.6) | 110 (22.0) |
| 50 – 59 | 12.7 | 295 (19.7) | 69 (13.9) | 129 (25.8) | 97 (19.4) |
| > 60 | 12.1 | 206 (13.8) | 72 (14.5) | 76 (15.2) | 58 (11.6) |
| **Education Level[11]** |  |  |  |  |  |
| No education | 30.4 | 279 (18.7) | 173 (35.0) | 93 (18.6) | 13 (2.60) |
| Primary school | 27.5 | 678 (45.4) | 205 (41.4) | 205 (41.0) | 268 (53.6) |
| Junior High School | 16 | 229 (15.3) | 47 (9.50) | 97 (19.4) | 85 (17.0) |
| Senior High School | 18.6 | 210 (14.1) | 53 (10.7) | 83 (16.6) | 74 (14.8) |
| Diploma or above | 7.6 | 99 (6.60) | 17 (3.40) | 22 (4.40) | 60 (12.0) |
| **Socio-Economic Status[44]** |  |  |  |  |  |
| Poor | 89.6 | 449 (30.0) | 151 (30.5) | 105 (21.0) | 193 (38.6) |
| Average | 4.80 | 860 (57.5) | 286 (57.8) | 331 (66.2) | 243 (48.6) |
| Rich | 5.70 | 186 (12.4) | 58 (11.7) | 64 (12.8) | 64 (12.8) |

<sup>a</sup> The percentage of people in different age groups at national level has been calculated based on the total people of aged above 15 years; <sup>b</sup> High: East Sumba District, Moderate: Belu District and low: East Manggarai District.

### Major malaria knowledge by malaria endemic settings in the ENTP

The difference of malaria knowledge on various aspects amongst three different settings is shown in Table 2. In terms of basic malaria understanding, the percentage of respondents hearing malaria terms and being aware that malaria could be prevented was high accounting for 86.1% with 95% confidence interval (CI): 84.2 – 88.0, p < 0.001 and 81.3%, 95% CI: 79.1 – 83.5, p < 0.001 respectively, whilst the awareness of malaria has dangerous effect on health was only 64.1%, 95% CI: 61.1 – 67.1, p < 0.001 with the highest in low MES (73.4%, 95% CI: 68.9 – 77.9, p < 0.001)) and the lowest in moderate MES (45.8%, 95% CI : 39.3 – 52.3).

**Table 2.**
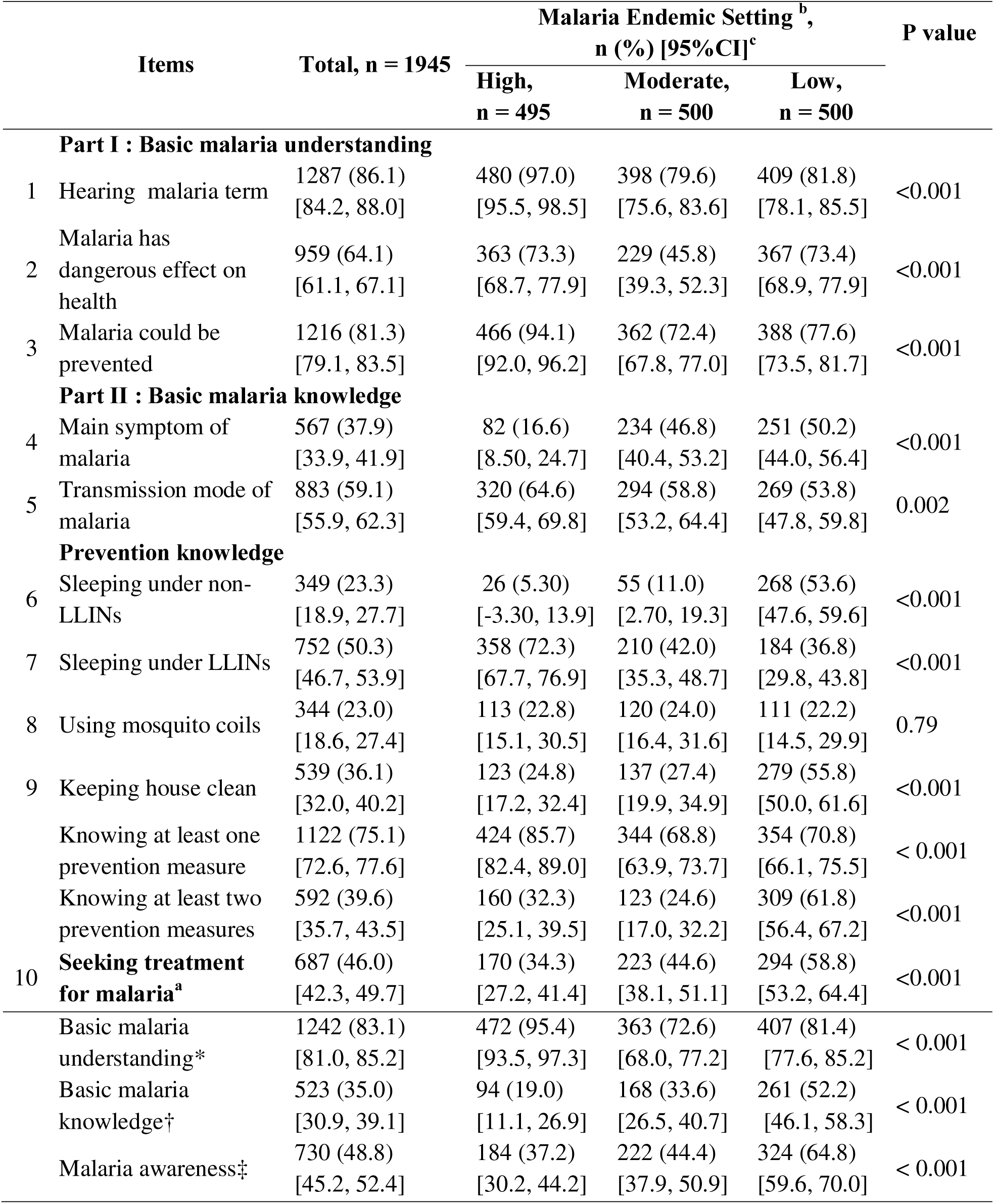
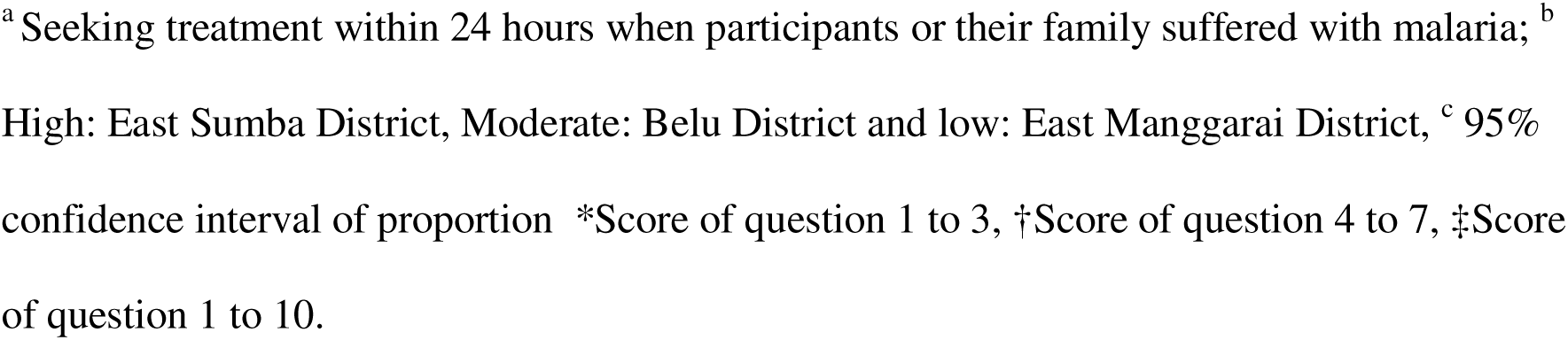
Distribution of major malaria knowledge of rural adults in three different malaria endemic settings (MES) in the East Nusa Tenggara Province (ENTP), Indonesia.

| Items |  | Total, n = 1945 | Malaria Endemic Setting <sup>b</sup>,<br>n (%) [95%CI]<sup>c</sup> |  |  | P value |
| --- | --- | --- | --- | --- | --- | --- |
|  |  |  | High,<br>n = 495 | Moderate,<br>n = 500 | Low,<br>n = 500 |  |
| Part I : Basic malaria understanding |  |  |  |  |  |  |
| 1 | Hearing malaria term | 1287 (86.1)<br>[84.2, 88.0] | 480 (97.0)<br>[95.5, 98.5] | 398 (79.6)<br>[75.6, 83.6] | 409 (81.8)<br>[78.1, 85.5] | <0.001 |
| 2 | Malaria has dangerous effect on health | 959 (64.1)<br>[61.1, 67.1] | 363 (73.3)<br>[68.7, 77.9] | 229 (45.8)<br>[39.3, 52.3] | 367 (73.4)<br>[68.9, 77.9] | <0.001 |
| 3 | Malaria could be prevented | 1216 (81.3)<br>[79.1, 83.5] | 466 (94.1)<br>[92.0, 96.2] | 362 (72.4)<br>[67.8, 77.0] | 388 (77.6)<br>[73.5, 81.7] | <0.001 |
| Part II : Basic malaria knowledge |  |  |  |  |  |  |
| 4 | Main symptom of malaria | 567 (37.9)<br>[33.9, 41.9] | 82 (16.6)<br>[8.50, 24.7] | 234 (46.8)<br>[40.4, 53.2] | 251 (50.2)<br>[44.0, 56.4] | <0.001 |
| 5 | Transmission mode of malaria | 883 (59.1)<br>[55.9, 62.3] | 320 (64.6)<br>[59.4, 69.8] | 294 (58.8)<br>[53.2, 64.4] | 269 (53.8)<br>[47.8, 59.8] | 0.002 |
| Prevention knowledge |  |  |  |  |  |  |
| 6 | Sleeping under non-LLINs | 349 (23.3)<br>[18.9, 27.7] | 26 (5.30)<br>[-3.30, 13.9] | 55 (11.0)<br>[2.70, 19.3] | 268 (53.6)<br>[47.6, 59.6] | <0.001 |
| 7 | Sleeping under LLINs | 752 (50.3)<br>[46.7, 53.9] | 358 (72.3)<br>[67.7, 76.9] | 210 (42.0)<br>[35.3, 48.7] | 184 (36.8)<br>[29.8, 43.8] | <0.001 |
| 8 | Using mosquito coils | 344 (23.0)<br>[18.6, 27.4] | 113 (22.8)<br>[15.1, 30.5] | 120 (24.0)<br>[16.4, 31.6] | 111 (22.2)<br>[14.5, 29.9] | 0.79 |
| 9 | Keeping house clean | 539 (36.1)<br>[32.0, 40.2] | 123 (24.8)<br>[17.2, 32.4] | 137 (27.4)<br>[19.9, 34.9] | 279 (55.8)<br>[50.0, 61.6] | <0.001 |
|  | Knowing at least one prevention measure | 1122 (75.1)<br>[72.6, 77.6] | 424 (85.7)<br>[82.4, 89.0] | 344 (68.8)<br>[63.9, 73.7] | 354 (70.8)<br>[66.1, 75.5] | < 0.001 |
|  | Knowing at least two prevention measures | 592 (39.6)<br>[35.7, 43.5] | 160 (32.3)<br>[25.1, 39.5] | 123 (24.6)<br>[17.0, 32.2] | 309 (61.8)<br>[56.4, 67.2] | <0.001 |
| 10 | Seeking treatment for malaria<sup>a</sup> | 687 (46.0)<br>[42.3, 49.7] | 170 (34.3)<br>[27.2, 41.4] | 223 (44.6)<br>[38.1, 51.1] | 294 (58.8)<br>[53.2, 64.4] | <0.001 |
|  | Basic malaria understanding* | 1242 (83.1)<br>[81.0, 85.2] | 472 (95.4)<br>[93.5, 97.3] | 363 (72.6)<br>[68.0, 77.2] | 407 (81.4)<br>[77.6, 85.2] | < 0.001 |
|  | Basic malaria knowledge† | 523 (35.0)<br>[30.9, 39.1] | 94 (19.0)<br>[11.1, 26.9] | 168 (33.6)<br>[26.5, 40.7] | 261 (52.2)<br>[46.1, 58.3] | < 0.001 |
|  | Malaria awareness‡ | 730 (48.8)<br>[45.2, 52.4] | 184 (37.2)<br>[30.2, 44.2] | 222 (44.4)<br>[37.9, 50.9] | 324 (64.8)<br>[59.6, 70.0] | < 0.001 |

<sup>a</sup> Seeking treatment within 24 hours when participants or their family suffered with malaria; <sup>b</sup> High: East Sumba District, Moderate: Belu District and low: East Manggarai District, <sup>c</sup> 95% confidence interval of proportion \*Score of question 1 to 3, †Score of question 4 to 7, ‡Score of question 1 to 10.

In terms of basic malaria knowledge, the awareness of fever as the main symptom of malaria was low (37.9%, 95% CI: 33.9 – 41.9, p < 0.001), which was 50.2%, 95% CI: 44.0 – 56.4, p < 0.001 in the low MES, 46.8%, 95% CI : 40.4 – 53.2, p < 0.001 in the moderate and 16.6%, 95% CI : 8.50 – 24.7, p < 0.001 in the high malaria endemic settings (P<0.001).The awareness of mosquito bites as the main cause of malaria was also low (59.1%, 95% CI: 55.9 – 62.3, p < 0.002), the highest in high MES (64.6%, 95% CI: 59.4 – 69.8, p < 0.002) and the lowest in low MES (53.8%, 95% CI: 47.8 – 59.8, p < 0.002)

The percentage of participants knowing at least one prevention measure to prevent malaria was high accounting for 75.1%, 95% CI: 72.6 –77.6, p < 0.001 with 85.7%, 95% CI: 82.4 – 89.0, p < 0.001 in high malaria endemic settings followed by 70.8%, 95% CI: 66.1 –75.5, p < 0.001 in low malaria endemic settings and 68.8%, 95% CI: 63.9 – 73.7, p < 0.001 in moderate malaria endemic settings (P<0.001). Whilst, the proportion of participants knowing at least two malaria prevention measures was low, which is only 39.6%, 95% CI: 35.7 – 43.5, p < 0.001 the highest 61.8%, 95% CI: 56.4 – 67.2, p < 0.001 in low endemic settings, followed by 32.3%, 95% CI: 25.1 – 39.5, p < 0.001 in high settings and 24.6%, 95% CI: 17.0 –32.2, p < 0.001 in moderate settings (P<0.001). The percentage of participants knew that sleeping under LLINs to prevent malaria was also low, which is only 50.3%, 95% CI: 46.7 – 53.9, p < 0.001, the highest 72.3%, 95% CI: 67.7 – 76.9, p < 0.001 in high MES, followed by 42%, 95% CI: 35.3 – 48.7, p < 0.001 in moderate and 36.8%, 95% CI: 29.8 – 43.8, p < 0.001 in low MES.

In terms of malaria treatment seeking behaviour, the proportion of participants who were awareness to seek treatment within 24 hour if they or their family members suffered with malaria was also low (46%, 95% CI: 42.3 – 49.7, p < 0.001), the highest 58.8%, 95% CI: 53.2 – 64.4, p < 0.001 in the low MES, 44.6%, 95% CI: 38.1 – 51.1, p < 0.001 in the moderate MES and 34.3%, 95% CI: 27.2 – 41.4, p < 0.001 in the high MES and the level of awareness was significantly different (P<0.001).

Overall, only 48.8% of rural adults in ENTP had malaria knowledge scores above 60% correct and 17.4% had above 80% correct. The proportion of participants having poor malaria knowledge score was high accounting for 42.9%, with 60.4% in high endemic settings followed by 44.2% in moderate endemic settings and 24.4% in low endemic settings as shown in Fig 2.

**Fig 2.**
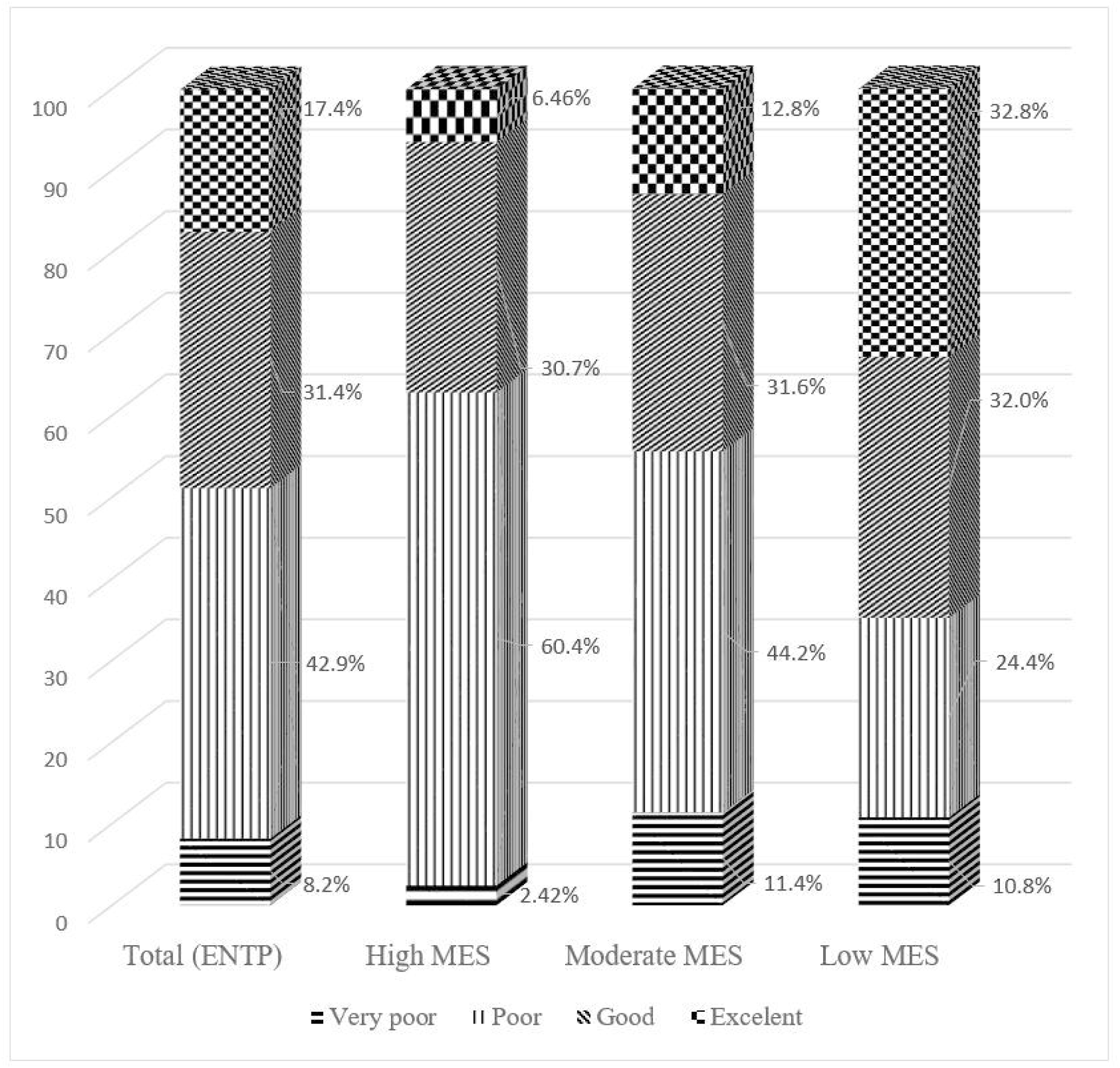
Distribution of malaria knowledge score amongst participants.

### Malaria awareness of rural adults in the ENTP

Among the participants, the percentage of awareness in basic malaria understanding was very high accounting for 83.1% (95% confidence interval (CI): 81.0 – 85.2, p < 0.001) with 95.4% (95%CI: 93.5 – 97.3, p < 0.001) in high malaria endemic settings followed by 72.6% (95% CI: 68.0 – 77.2, p < 0.001) in moderate malaria endemic settings and 81.4% (95% CI: 77.6 – 85.2 in low malaria endemic settings (P<0.001). The proportion of rural adults having the awareness of basic malaria knowledge was low, which was only 35% (95% CI: 30.9–39.1, p < 0.001), the highest 52.2% (95% CI: 46.1 – 58.3, p < 0.001) in the low API settings, followed by 33.6% (95% CI: 26.5 – 40.7, p < 0.001) and 19.0% (95% CI: 11.1– 26.9, p < 0.001) in the moderate and high MES settings, respectively.

Overall, only 48.8% (95% CI: 45.2 – 52.4, p < 0.001) of rural adults in the ENTP had malaria awareness. The malaria awareness in low MES was the highest (64.8%, 95% CI: 59.6–70.0, p < 0.001) followed by 44.4% (95% CI: 37.9 – 50.9, p < 0.001) in the moderate malaria endemic settings and 37.2% (95% CI: 30.2 – 44.2, p < 0.001) in the high malaria endemic settings. The difference in awareness was statistically significant amongst these three settings (P<0.001) as shown in Table 2.

From Table 3, it shows that the highest proportion of participants having basic malaria understanding was in high endemic setting (95.4%), meanwhile the highest proportion of participants having basic malaria knowledge and malaria awareness was in low endemic settings, accounting for 52.2% and 64.8% respectively. The basic malaria knowledge for participants living in low malaria endemic setting was almost five times higher than that of in high endemic setting (Odds ratio (OR): 4.66, 95% confidence interval (CI): 3.50– 6.20, p < 0.001). Rural adults residing in low endemic settings were associated with 311% higher prevalence of malaria awareness compared to rural adults in high endemic settings (OR: 3.11, 95% CI: 2.40 – 4.03, p < 0.001).

**Table 3.**
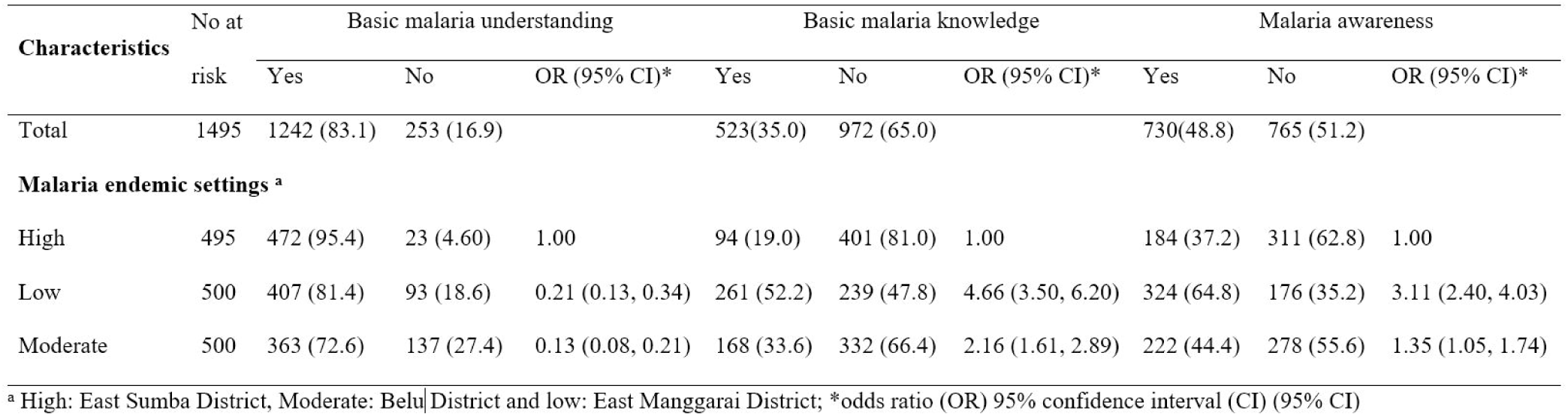
The Odds ratio of basic malaria understanding, basic malaria knowledge and malaria awareness amongst three types of malaria endemic settings in East Nusa Tenggara Province, Indonesia.

## Discussion

This is the first population-based study focusing on malaria awareness of rural adults amongst three malaria endemic settings from ENTP since the launch of national commitment of Indonesia government to eliminate malaria by 2030. The main finding of the study was that malaria awareness of rural adults was very low, which causes a significant barrier to malaria elimination in the region. The results indicate that malaria awareness of rural adults in high MES was the lowest of the other MES.

This study shows that a high proportion of rural adults in high and moderate MES has poor malaria knowledge. This finding was consistent with another study in Southern Africa [45] that revealed that residents in high MES had lower malaria knowledge compared with those in low MES. However, this finding was contrast with study in China [37], Bangladesh [22], Eritrea [46], North Sudan [47] and India [48] revealing that high malaria knowledge for rural population in high MES. This discrepancy might be explained with the fact that the rural community in those countries had been exposed with various interventions to improve malaria knowledge [22, 37, 38, 46, 47, 49] and in China, the government had included malaria awareness index as one of the action plan for malaria elimination since 2010 [50]. Meanwhile, in ENTP the interventions to improve malaria awareness of rural community was not documented yet. Findings of the study indicates that more attention should be paid for rural adults in high and moderate MES to escalate the malaria elimination. However, much attention should be paid to rural adults in low MES considering that high numbers of inter province migration flow [51] and inter district migration flow [52] could lead to imported malaria cases in this province.

Findings of this study also indicated that basic malaria knowledge of rural adults was very low. Only about 38% of rural adults could identify fever as the main symptom of malaria. This finding indicates that more than half of rural adults did not have the ability to identify correctly the main symptom of the disease which may lead to low level of awareness for malaria infection. These result contrasted a study conducted in Cabo Verde [39], that was on track to achieve malaria elimination zone by 2020 [3], and in Iran [40]which indicated that the high level of participants could identify fever as the main symptom of malaria. With regard to transmission mode of malaria, more than half of rural adults knew that malaria was caused by mosquito bite. This proportion is lower than reported in other countries [39, 40, 46, 48, 53] which revealed that the majority of rural communities recognised mosquito bites as the main cause of malaria. The finding of this study indicated that there is still a large proportion of rural adults with a lack of awareness for protecting themselves from mosquito bites. Failure to improve the awareness of this community leads to low levels of the usage of malaria prevention methods promoted by the Indonesia government which in turn will increase the burden of malaria in this province.

There are four malaria prevention measures that are familiar to the rural adults in the ENTP, including sleeping under non-LLINs, using mosquito coils, keeping houses clean and sleeping under LLINs. However, the proportion of participants that knew these methods was very low and disparity amongst MES for this knowledge was marked. It is worth noting that the percentage of rural adults having knowledge at least one prevention measure is high whereas the proportion of rural adults having knowledge of at least two prevention methods was very low. Combining various methods to prevent malaria is more effective than only one approach [54].

In our study, the proportion of rural adults having knowledge of sleeping under LLINs to prevent malaria was low. This finding contrasts with studies in other countries such as Tanzania [55], Eritrea [46], North Sudan [47], Iran [56], Bangladesh [57] and Southern Africa [45] which revealed a high proportion of rural community knew that sleeping under treated nets was a protective method to prevent malaria. This disparity might be because of different levels of knowledge in transmission mode of malaria. In those countries, most of the rural population could identify correctly the main cause of malaria, whilst, in this research only about half of the studied population knew malaria was caused by mosquito bite. Failure to improve the awareness of communities about the benefits of sleeping under LLINs provided a negative impact on the malaria elimination program. Systematic review on the use of LLINs indicated that despite LLINs being provided free of charge and supported by government agencies and many non-government organisations, lack of awareness among communities lead them to misuse of LLINs, for instance for protecting and storage of food material [58].

The study revealed that there is a significant difference regarding knowledge of sleeping under LLINs amongst three different settings. The highest percentage for this knowledge was for rural adults in high endemic settings, followed by moderate settings and the lowest was in low settings. This finding is consistent with study in Bangladesh [22] and Colombia [59]. The high level of this knowledge in high endemic settings might be due to long term exposure with LLINs distribution program in the region. It is understood that in malaria endemic communities with many ongoing programs of malaria intervention, the level of malaria prevention knowledge should be higher compared to other areas which have less malaria prevention programs. Since 2008, the target program of LLINs distribution in East Sumba district was higher compared with other districts [14] and in 2017 during the mass campaign of LLINs in the country, this district was also included in the program [15].

Regarding perceptions on treatment seeking behaviour, our study found that the awareness of seeking treatment within 24 hours when participants or their family members suffer with malaria was poor. This is consistent with other studies in some parts of Indonesia [24, 60], and other South East Asia countries such as Myanmar [21], India [61], Bangladesh [57] and Cambodia [62]. The poor level of seeking malaria treatment in this study might occur since over one third of the total participants still believed that malaria was not dangerous for their health. Therefore, they treated malaria at home first for several days before they visited a local health centre. Prompt treatment seeking behaviour is critical to escalate for malaria elimination. With considering low awareness of rural adults in the ENTP, more efforts are needed to improve the awareness of the rural adults since failure to seek treatment within 24 hours after onset of the clinical symptoms lead to increased fatality rate [63].

Community engagement is fundamental to malaria elimination [64]. To improve the community participation, the community awareness should be measurable. The study indicates that malaria awareness of participants is poor and malaria awareness index is not part of the malaria elimination program of the ENTP currently [65], therefore, malaria awareness index should be part of the key strategic interventions of the ENTP to improve malaria awareness of the community. Having this index in their malaria elimination program enables the local authority for implementing and evaluating the progress of malaria awareness of the local community at district, sub-district and village level. Furthermore, the improvement of awareness on infectious disease, including malaria, enables community to improve their self-protection and seek early treatment [66], finding treatment source preference [67], reduce the malaria prevalence [22], and finally it could boost malaria elimination [68].

It is suggested the partnership between the health department and education department of the ENTP plays a significant role in promoting malaria knowledge as a part of local curriculum to improve malaria awareness index of local community. Students could be an important agent for change. They could be encouraged to share their malaria knowledge with their family as be shown in other countries [69, 70]. The great achievement of the Chinese government to achieve zero local malaria transmission for the first time in 2017 was also supported by the massive effort to improve malaria awareness of communities including school children [71]. Considering that a high proportion of residents in rural areas in the ENTP have no education level [11, 72], the malaria education program in countryside schools could improve malaria awareness of rural communities.

This research is the first reliable data on malaria awareness and knowledge in the general population in ENTP Indonesia, particularly for adults living in remote areas. Data provides a large and representative sample size for this population. The potential weakness of our study is that data collection had been collected in one time period and from one province. The study needs to be repeated in a random sample in other regions for capturing a truly representative sample of rural adults nationally. Because of limited resources for the study, we cannot check the inter or intra interviewer’s reliability. The interviewers do not have a chance to interview the same research participants, indicating that inter-intra reliability could not be evaluated. However, interviewers were selected for those having certified degree in nursing and they participated in one day intensive training applying the same approach. Despite those limitations, findings of the study have provided insights into the level of malaria knowledge of rural adults of the ENTP.

## Conclusions

Malaria awareness of rural adults needs to be improved. Public health programs of the local government should incorporate malaria awareness index as a key intervention and this index should be measurable by setting up the reasonable target to improve the awareness of the local communities. Having this index in malaria elimination programs of the ENTP will help local authorities to manage and evaluate the progress of malaria awareness of the local community at district, sub-district and village level. Public health campaigns should be focused on improving basic malaria knowledge such as main symptom, transmission mode, prevention methods of malaria, seeking early treatment behaviour for rural adults in the province. This method will support a national action plan for malaria elimination in the country. Failure to address the awareness in rural communities will mean malaria elimination will fall short.

## Data Availability

Data will be available after the article has been accepted to publish.

## Acknowledgments

We thank all respondents for their participation in this project. Our gratitude also dedicated to the governor of ENTP, head of East Sumba, Belu, and East Manggarai District, nine head of sub-districts, and forty-nine village leaders for allowing conducted this research in their region.

## Authors’ Contributions

RDG designed the study, prepared the data collection instruments, organised ethics, conducted primary data collection, analysed the data and wrote the draft manuscript. FMAI and JK supervised the study, reviewed the paper and provided substantial inputs. All authors have approved the manuscript for submission.

## Supporting Information

S1 Fig 1: Map of Study Sites

S2 Fig 2. Distribution of malaria knowledge score amongst participants.

S1 Table 3: The Odds ratio of basic malaria understanding, basic malaria knowledge and malaria awareness amongst three types of malaria endemic settings in East Nusa Tenggara Province, Indonesia.

S1 Data: Database for study malaria awareness in East Nusa Tenggara Province Indonesia

